# Agreeability testing of AMSTAR-PF, a tool for quality appraisal of systematic reviews of prognostic factor studies

**DOI:** 10.1101/2025.04.10.25325555

**Authors:** ML Henry, NE O’Connell, RD Riley, KGM Moons, BJ Shea, L Hooft, SB Wallwork, JAA Damen, N Skoetz, RP Appiah, C Berryman, SM Crouch, GA Ferencz, AR Grant, KM Henry, AM Herman, EL Karran, I Koralegedera, HB Leake, E MacIntyre, B Mouatt, K Phuentsho, DA Van Der Laan, E Welsby, LK Wiles, EM Wilkinson, MK Wilson, MV Wilson, GL Moseley

**Author notes:** Corresponding Author N.E. O’Connell, PhD, Centre for Health and Wellbeing Across the Lifecourse, Department of Health Sciences, Brunel University London, Uxbridge UB8 3PH, United Kingdom.

## Abstract

**Background:** This paper details initial testing of the agreeability and usability of a novel quality appraisal tool for systematic reviews of prognostic factor studies: AMSTAR-PF.

**Methods:** Fourteen appraisers each assessed eight systematic reviews using AMSTAR-PF. Their ratings for each question and each article were compared, with interrater, inter-pair and intrapair agreeability calculated using Gwet’s agreement coefficient. Time of use and time to reach consensus were also recorded.

**Results:** Interrater agreement averaged 0.59 (range, 0.21-0.90), inter-pair 0.61 (range 0.24-0.91) and intrapair 0.75 (range 0.45-0.95) across the domains, with agreement for the overall rating 0.46 (95%CI 0.30-0.62) for interrater, 0.46 (95%CI 0.17-0.74) for inter-pair, and 0.68 (range of averages 0.22-1.00) for intrapair agreement. The majority (60.7%) of intrapair ratings were identical, with 94.6% of final ratings either identical or only one category different for the overall appraisal. The time taken to appraise a study with AMSTAR-PF improved with use and averaged around 34 minutes after the first two appraisals.

**Conclusions:** Despite some variance in agreeability for different domains and between different appraisers, the testing results suggest that AMSTAR-PF has clear utility for appraising the quality of systematic reviews of prognostic factor studies.

## Introduction

There is an increasing number of studies and systematic reviews investigating prognostic factors (PFs)(1, 2). PFs are variables associated with a certain outcome (3), and are particularly useful in providing patients with a prognosis and developing prognostic models (eg, for outcome risk prediction). PFs can include patient demographics, such as age and sex; clinical signs and symptoms, such as imaging results and severity of symptoms; and broader environmental factors such as location of residence. Published studies of prognosis research are variable in quality, and subject to a range of potential biases that can impact study findings and decrease confidence in the results (1, 4). As the body of primary prognosis research grows, so will the demand for high quality evidence syntheses. Until recently, there has been no standardised tool specifically developed to appraise the quality of reviews of prognostic factor studies. In order to fill this gap, our group developed A MeaSurement Tool to Assess systematic Reviews of Prognostic Factor studies (AMSTAR-PF) (5), through a multi-stage process, by a team including people with expertise in prognostic factor reviews, evidence synthesis and quality appraisal tool development.

AMSTAR-PF consists of 14 domains, some of which have subsections, resulting in 19 specific questions that inform an overall judgement on the quality of the review (see Appendix I in Supplementary material). All 19 questions have answer options of Yes (Y), Probably Yes (PY), Probably No (PN), and No (N). Six questions also have an option of Not Applicable (N/A). The final judgement has four options for the overall quality of the review: High, Moderate, Low, and Critically Low. As part of the development process, we sought to test AMSTAR-PF to determine measures of agreement and usability.

Agreement relates to the reliability of a tool and has two main subdivisions; how consistently a given item is rated by the same rater (or same group of raters) on different occasions, and how similarly two or more different raters (or groups) rate the same item, which is the agreement we were interested in for this paper. Usability of a tool can consider reported difficulties or ease of use, as well as objective measures such as time to use. In this paper, we aimed to determine agreement and usability of AMSTAR-PF, using (i) measures of interrater, inter-pair, and intrapair agreement, and (ii) time to complete appraisal using the tool by researchers with a diverse range of research experience.

## Methods

We pre-registered our protocol on OSF prior to testing (osf.io/acrwf), and received ethics approval (ID: 206951) from the University of South Australia’s Human Research Ethics Committee.

The testing used a convenience sample of eight systematic reviews of prognostic factor studies. Each systematic review will herein be referred to as an ‘article’. The selected articles included two in low back pain (6, 7), two in cancer (8, 9), two in brain injury (10, 11), and two in COVID-19 (12, 13), and included meta-analyses and narrative syntheses. Two articles (6, 9) were Cochrane reviews. The articles were selected to cover a range of different clinical areas, though the choice of the four areas was somewhat arbitrary. Back pain and concussion were topics of an umbrella review being undertaken by some of the research group, cancer research was deemed likely to have a selection of prognostic factor reviews, and COVID-19, being a newly emerged disease, was chosen to represent current practice in prognostic factor research. The two Cochrane reviews were selected from the Cochrane Library website, with the prognosis filter applied, to match one of the four topics. The remaining articles were selected based on their titles from a Google Scholar search, and ensuring a mix of reviews with and without meta-analyses.

Testing of the tool was completed by appraisers who were independent of the process of developing AMSTAR-PF, and who had not been involved in any of the preliminary pilot testing stages. Appraisers were recruited via an email sent to 43 people associated with a university research group. We limited the number of appraisers to 14 for pragmatic reasons. Each of the eight articles was appraised by the 14 appraisers (seven pairs). Where possible, pairs consisted of researchers with different experience levels (eg, a student with a post-doctoral researcher), because we considered that this would most faithfully reflect the likely use of the tool in research practice.

## Agreement

All appraisers were provided with the AMSTAR-PF tool and Guidance Notes (5) and asked to familiarise themselves with these documents prior to assessing the articles. Appraisers had an opportunity to ask questions and seek further clarification from the lead author (MLH), if needed. Each appraiser then independently applied the AMSTAR-PF tool on two articles – one Cochrane review article and one non-Cochrane article. Appraiser pairs met to discuss their appraisal, reach consensus on these first two articles, discuss consistency in the application of the tool and develop any decision-making rules they would implement, as a pair, for the remainder of the articles. This practice is recommended (14) and often used when undertaking quality appraisals of articles, and reflects how the tool is intended to be used. Once the pair was satisfied with their consensus, each appraiser independently appraised the remaining six articles. When both appraisers in a pair had completed the appraisals for the remaining six articles, they again compared findings and reached consensus for these six articles. For all eight articles, the overall rating for each article as well as the responses to each question within AMSTAR-PF were compared; agreements and discrepancies were recorded.

## Timing

For each of the articles, appraisers were asked to read the article first and then record the time from the start of the AMSTAR-PF appraisal process to completion. Each appraiser pair also recorded the total time taken to reach consensus for each of the articles, which consisted of consensus on the overall rating as well as each of AMSTAR-PF’s 19 questions. Appraisers were asked to record and describe any instances where they could not reach consensus.

To mitigate any effect of order on ratings, agreeability, or timing, we counterbalanced the order of the eight articles across appraiser pairs, with three pairs randomised to complete one sequence (Aldin et al. (9), Izcovich et al. (12), Maglietta et al. (13), Mercier et al. (11), Pinheiro et al. (7), Puig et al. (10), Wijnands et al. (8), Hayden et al. (6)), and four pairs completing the reverse sequence.

## Analysis

Interrater agreement (the agreement across all 14 appraisers), inter-pair agreement (the agreement across the consensus scores of the seven pairs) and intrapair agreement (the mean of the agreement between the two members of each of the seven pairs) were calculated across the eight articles, both overall and for each of the individual questions. For the domain questions, we calculated agreement with the original answering options (Yes (Y), Probably Yes (PY), Probably No (PN), and No (N), and for some questions, N/A), as well as with the Y/PY, and N/PN answers collapsed, as per our protocol.

We used Gwet’s Agreement Coefficient (AC) in our analysis of the agreement. This was a deviation from our original protocol, which stipulated using Cohen’s and Fleiss’ Kappa for analyses. We made this deviation after data collection but prior to commencing any analysis, after receiving statistical advice. Gwet’s AC is a chance-corrected agreement coefficient that was developed in part due to instability of Kappa statistics across a range of agreement levels, in particular paradoxes in which a high level of agreement can lead to low Kappa scores (15-17). Gwet’s AC can be used with multiple raters and has been shown to provide stable measures of agreement across a range of agreement levels. Gwet’s AC1 was calculated for nominal data (questions 2b, 7c, 9a, 9b, 10, and 12, which include an N/A option), while Gwet’s AC2, with a linear weighting, was used for the ordinal data (the remaining 13 questions and the overall judgement) (18). Consistent with our original protocol, we have also calculated Cohen’s and Fleiss’ Kappa, which can be found in Appendix IV of the supplementary material. Calculations were performed within Stata/SE 18.0 (StataCorp LLC).

Interpretations of the agreement coefficients were based on Landis and Koch’s benchmarks (19) and calculated in the manner recommended by Gwet (18). This manner considers the standard error to give a probability of the value falling within a certain benchmark category. We used the recommended 95% cumulative probability as the cut-off level (18). RStudio version 2024.04.1 Build 748 (RStudio Inc) and the irrCAC package were used for calculations, and ggplot or Microsoft Excel (version 2408, Microsoft Corporation) for visualisation. The category labels for Gwet’s AC were aligned with Landis and Koch’s suggestions: < 0, Poor; 0.0-0.2, Slight; 0.2-0.4, Fair; 0.4-0.6, Moderate; 0.6-0.8, Substantial; and 0.8-1.0, Almost Perfect (19).

We also presented the raw numbers of agreements and disagreements for each of the intrapair decisions. This was a deviation from protocol, but we believe adds interpretability and clarity.

Descriptive statistics were calculated for ‘time taken to use the tool’, and ‘time taken to reach consensus’.

To detect an effect of the consensus process or learning on level of agreement, we compared agreement for the first two articles appraised to agreement for the next six, and agreement for the first two articles to agreement for the final two. We also investigated whether there were differences in agreement between the two different orders of articles. We did this by comparing Gwet’s AC for the two orders, and by comparing the final ratings of each article. Furthermore, we compared agreement for the Cochrane articles to those for the non-Cochrane articles. The timing of completion was compared for the order of completion, as well as for the experienced (seven researchers with five or more years in research) versus novice (seven researchers with three or fewer years in research) researchers within the 14 appraisers.

To detect group differences in agreement and timings, we used paired t-tests when data were normally distributed, and Wilcoxon Signed Rank or Rank Sum tests when they were not. A significance level of p < 0.05 was used.

## Results

### Demographics of appraisers

The 14 appraisers (11 female, 3 male) were aged between 22-59 years old and had experience in research ranging from less than one year to over 10 years. Appraisers included Honours, Masters, and PhD students, postdoctoral researchers, and research fellows. Eleven spoke English as their first language and three as an additional language. One appraiser had previous experience with prognostic factor research. Twelve had experience with pain research. None had research experience with any of the other three topics addressed in the articles (COVID-19, brain injury, and cancer). Twelve had experience of performing risk of bias and/or quality appraisal assessments, including three who had used AMSTAR/AMSTAR 2.

### Interrater, inter-pair and intrapair agreement across the domains of the AMSTAR-PF

Interrater agreement across the 14 appraisers ranged between slight and substantial (Gwet’s AC mean 0.59, 95%CI 0.48-0.70, range 0.21 – 0.90) for the 19 questions of AMSTAR-PF, and inter-pair agreement also ranged between slight and substantial (mean 0.61, 95%CI 0.49-0.73, range 0.24 – 0.91). Intrapair agreement was taken as the mean of the seven intrapair AC scores, and ranged between fair to almost perfect agreement (mean 0.75, 95%CI 0.68-0.82, range of means 0.45 – 0.95). See Table 1 and Figure 1 for more detailed information. Of the 1064 domain intrapair decisions, 776 (72.9%) were identical, and 197 (18.5%) were one category different, meaning that 8.6% of intrapair decisions differed by more than one category, or involved one appraiser in a pair selecting the N/A option and the other member of the pair deeming the question applicable.

**Table 1.**
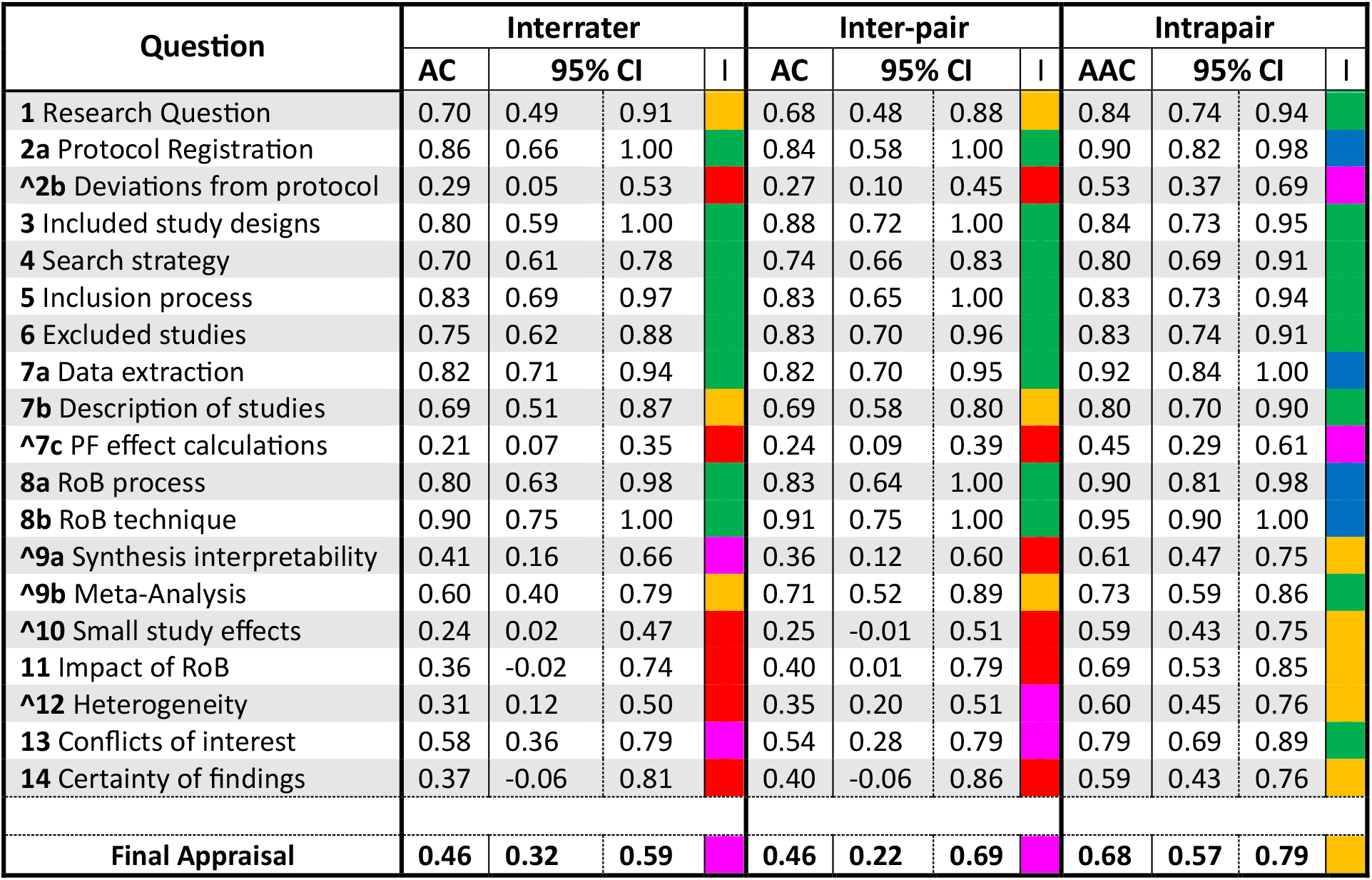
Gwet’s AC for each AMSTAR-PF question, with all answering options, and for the final judgement. Questions marked (⋀) had an N/A option and were treated as nominal data, calculated as Gwet’s AC1. All other questions were calculated as Gwet’s AC2 with linear weighting. Confidence intervals are capped at 1.00. Benchmark interpretation is calculated using 95% cumulative probabilities for Landis and Koch’s benchmark categories, 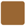 < 0, Poor; 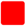 0.0-0.2, Slight; 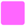 0.2-0.4, Fair; 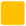 0.4-0.6, Moderate; 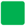 0.6-0.8, Substantial; and 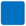 0.8-1.0, Almost Perfect. AC, Gwet’s Agreement Coeficient; Avg, average; I, Interpretation category; 95% CI, 95% Confidence Interval.

**Figure 1.**
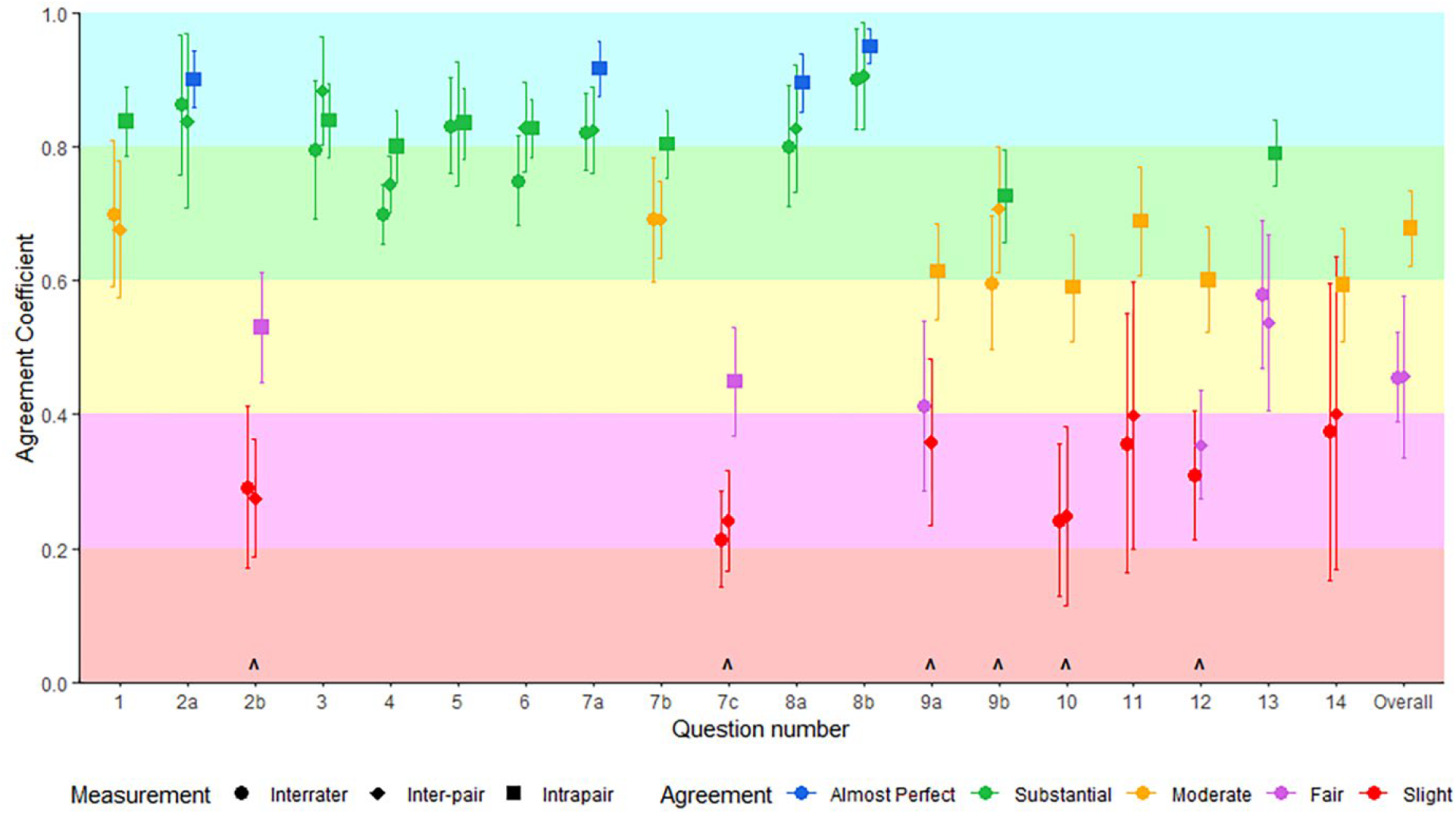
Gwet’s AC for each AMSTAR-PF question, with all answering options, and for the overall appraisal. Interrater and Inter-pair show AC and standard error as error bars. Intrapair show average AC across pairs with standard error of the mean (SEM) as the error bars. Questions marked (∧) had an N/A option and were treated as nominal data, calculated as Gwet’s AC1. All other questions were calculated as Gwet’s AC2 with linear weighting. Benchmark interpretation is colour-coded, with each plotted point coloured according to the calculated 95% cumulative probability for Landis and Koch’s benchmark categories; < 0, Poor; 0.0-0.2, Slight; 0.2-0.4, Fair; 0.4-0.6, Moderate; 0.6-0.8, Substantial; and 0.8-1.0, Almost Perfect.

When answering options were collapsed (from ‘Yes and Partial Yes’, and ‘No and Partial No’ to ‘Yes’ and ‘No’, and in some questions, N/A), the interrater agreement over the 19 questions ranged from slight to almost perfect (0.38 – 0.96), the inter-pair agreement ranged between poor and almost perfect (0.36 – 0.96), and the mean intrapair agreements were between moderate and almost perfect (0.63 – 0.97). See Table 2 and Figure 2. Of the 1064 intrapair decisions, 934 (87.8%) were the same direction, or both members of the pair agreed it to be N/A.

**Table 2.** Gwet’s AC1 for dichotomised answering responses (Y/PY, and N/PN collapsed) Questions marked (⋀) had an N/A option. Confidence intervals are capped at 1.00. Benchmark interpretation is calculated using 95% cumulative probabilities for Landis and Koch’s benchmark categories, 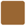 < 0, Poor; 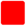 0.0-0.2, Slight; 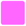 0.2-0.4, Fair; 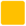 0.4-0.6, Moderate; 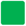 0.6-0.8, Substantial; and 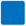 0.8-1.0, Almost Perfect. AC, Gwet’s Agreement Coefficient; Avg, average; I, Interpretation category; 95% CI, 95% Confidence Interval.

| Question | Interrater |  |  |  | Inter-pair |  |  |  | Intrapair |  |  |
| --- | --- | --- | --- | --- | --- | --- | --- | --- | --- | --- | --- |
|  | AC | 95% CI |  |  | AC | 95% CI |  |  | AAC | 95% CI |  |
| 1 Research Question | 0.81 | 0.60 | 1.00 |  | 0.87 | 0.66 | 1.00 |  | 0.91 | 0.80 | 1.00 |
| 2a Protocol Registration | 0.89 | 0.74 | 1.00 |  | 0.87 | 0.67 | 1.00 |  | 0.92 | 0.81 | 1.00 |
| ^2b Deviations from protocol | 0.40 | 0.13 | 0.68 |  | 0.49 | 0.18 | 0.80 |  | 0.63 | 0.47 | 0.79 |
| 3 Included study designs | 0.87 | 0.74 | 1.00 |  | 0.92 | 0.82 | 1.00 |  | 0.88 | 0.77 | 1.00 |
| 4 Search strategy | 0.85 | 0.71 | 0.99 |  | 0.92 | 0.82 | 1.00 |  | 0.86 | 0.74 | 0.99 |
| 5 Inclusion process | 0.89 | 0.79 | 0.99 |  | 0.94 | 0.80 | 1.00 |  | 0.87 | 0.75 | 0.99 |
| 6 Excluded studies | 0.92 | 0.79 | 1.00 |  | 0.94 | 0.82 | 1.00 |  | 0.97 | 0.90 | 1.00 |
| 7a Data extraction | 0.96 | 0.91 | 1.00 |  | 0.96 | 0.89 | 1.00 |  | 0.96 | 0.90 | 1.00 |
| 7b Description of studies | 0.77 | 0.54 | 1.00 |  | 0.63 | 0.26 | 1.00 |  | 0.83 | 0.68 | 0.97 |
| ^7c PF effect calculations | 0.44 | 0.16 | 0.71 |  | 0.50 | 0.25 | 0.75 |  | 0.64 | 0.47 | 0.80 |
| 8a RoB process | 0.90 | 0.73 | 1.00 |  | 0.92 | 0.73 | 1.00 |  | 0.96 | 0.90 | 1.00 |
| 8b RoB technique | 0.91 | 0.76 | 1.00 |  | 0.94 | 0.80 | 1.00 |  | 0.96 | 0.90 | 1.00 |
| ^9a Synthesis interpretability | 0.67 | 0.40 | 0.93 |  | 0.65 | 0.39 | 0.91 |  | 0.76 | 0.63 | 0.90 |
| ^9b Meta-Analysis | 0.86 | 0.71 | 1.00 |  | 0.96 | 0.86 | 1.00 |  | 0.88 | 0.78 | 0.98 |
| ^10 Small study effects | 0.41 | 0.15 | 0.67 |  | 0.36 | 0.08 | 0.63 |  | 0.76 | 0.61 | 0.90 |
| 11 Impact of RoB | 0.41 | -0.02 | 0.83 |  | 0.37 | -0.06 | 0.80 |  | 0.69 | 0.50 | 0.88 |
| ^12 Heterogeneity | 0.74 | 0.47 | 1.00 |  | 0.79 | 0.54 | 1.00 |  | 0.85 | 0.73 | 0.97 |
| 13 Conflicts of interest | 0.55 | 0.25 | 0.86 |  | 0.52 | 0.16 | 0.87 |  | 0.73 | 0.55 | 0.90 |
| 14 Certainty of findings | 0.38 | -0.06 | 0.82 |  | 0.36 | -0.12 | 0.85 |  | 0.63 | 0.43 | 0.84 |

**Figure 2.**
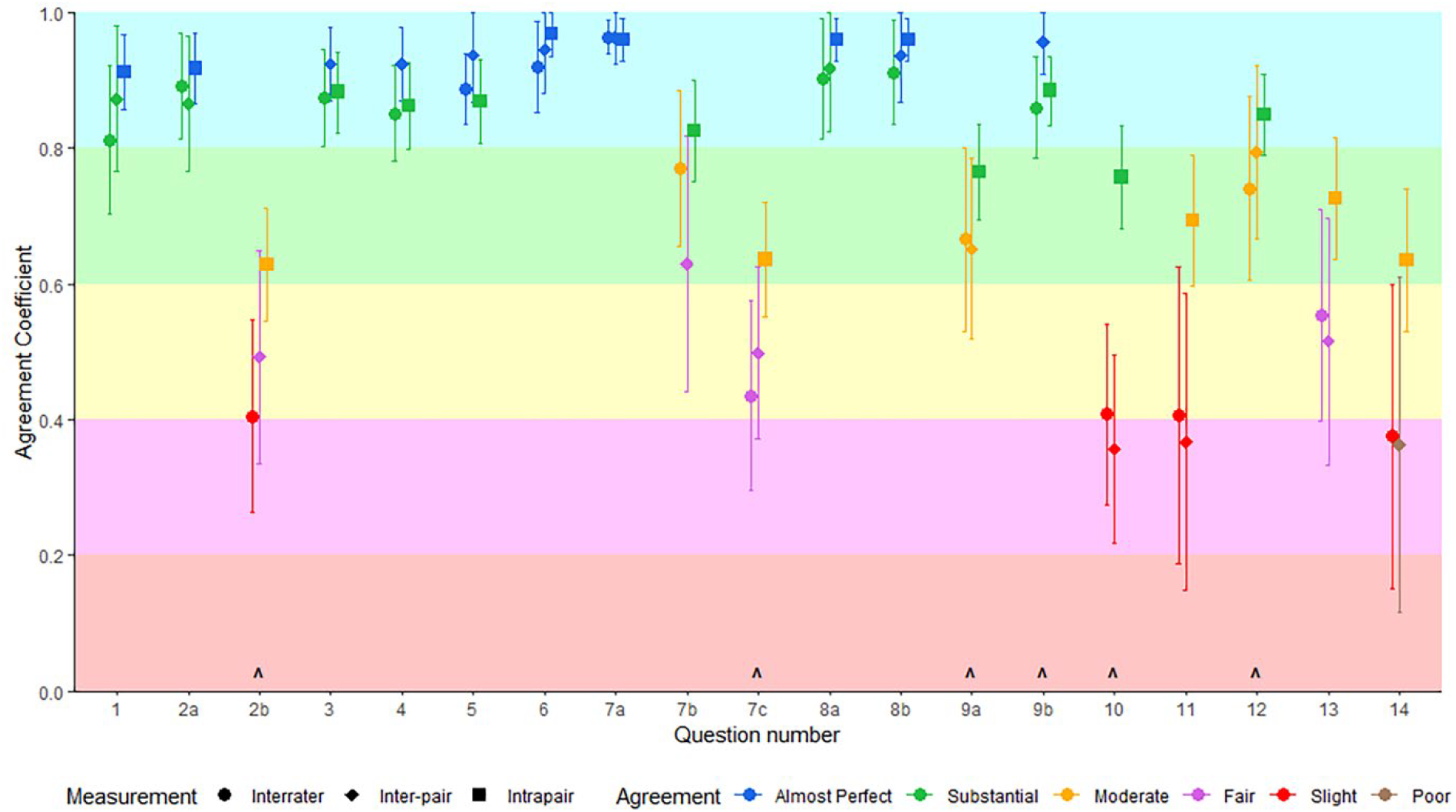
Gwet’s AC for each AMSTAR-PF question with dichotomised answering responses (Y/PY, and N/PN collapsed) Interrater and Inter-pair show AC and standard error as error bars. Intrapair show average AC across pairs with standard error of the mean (SEM) as the error bars. Error Bars are capped at 1.0. Questions marked (⋀) had an N/A option. Benchmark interpretation is colour-coded and calculated using 95% cumulative probabilities for Landis and Koch’s benchmark categories, < 0, Poor; 0.0-0.2, Slight; 0.2-0.4, Fair; 0.4-0.6, Moderate; 0.6-0.8, Substantial; and 0.8-1.0, Almost Perfect

Agreement scores tended to be higher for the earlier questions in AMSTAR-PF; questions that dealt with review planning, literature searching and study inclusion. Agreement for the latter questions regarding synthesis of results and analysis/interpretation of the included studies generally displayed lower levels of agreement.

### Interrater, inter-pair and intrapair agreement for the overall judgement using AMSTAR-PF

Gwet’s AC2 for 14 appraisers assessing the overall quality of the eight papers was fair 0.46 (95%CI 0.30-0.62) and the inter-pair agreement was also fair 0.46 (95%CI 0.17-0.74). We found moderate intrapair agreeability for overall quality across the seven pairs (average 0.68 (95%CI 0.44-0.91)).

Across the 56 intrapair agreements for overall quality, 34 (60.7%) were identical, and a further 19 (33.9%) were one category different. Three decisions (5.4%) differed by two categories.

### Subgroup Analysis

Agreement was similar for the first two articles appraised (mean (range of means) intrapair Gwet’s AC = 0.76 (0.42-1.00)) and the next six articles (0.74 (0.47-0.95)) (see Appendix II, Table A and Figure A, in the Supplementary material). We decided post-hoc to explore potential differences in agreement across the articles further, because we suspected there may be a difference in agreement with the Cochrane (the first and eighth articles appraised) and non-Cochrane articles, (see Appendix II, Table B and Figure B, in Supplementary material) because Cochrane articles are generally considered to be more rigorously developed and reported (20, 21). A difference in the proportion of Cochrane articles within the subgroups, therefore, may have influenced the average agreement and masked changes of agreement as appraisers proceeded. We found that agreement was significantly higher for the two Cochrane articles (mean intrapair AC 0.88 (range of means 0.67-1.00)) than it was for the non-Cochrane articles (0.71 (0.38-0.99)) (Wilcoxon Rank Sum test W = 317.5, p = 0.001). Both Cochrane articles were unanimously rated as high quality by every pair; none of the non-Cochrane articles received this level of agreement. Given this higher agreement in Cochrane articles, we performed an additional analysis comparing the first two articles appraised with the final two (because both combinations had one Cochrane and one non-Cochrane article). No statistically significant difference was found in average intrapair agreement (first two articles, (0.76 (range of means 0.42-1.00)); last two articles (0.83 (0.67-1.00)) (t(33.85) = −1.54, p = 0.133; (95%CI of difference −0.15 to 0.02)). An additional post-hoc analysis was performed to investigate the degree of intrapair difference in the final ratings given to the articles. By coding the final ratings (high, moderate, low, critically low) 1-4, respectively, we were able to assess for differences in the overall ratings given to articles. The first two articles averaged 0.50 categories different; the final two articles averaged 0.21 categories different, but this was not a statistically significant difference (Wilcoxon Signed Rank Test V = 27, p = 0.18).

Across all papers and domains, t-tests for interrater and inter-pair agreement did not suggest a difference between the orders of completion (t(37.74) = 0.54, p = 0.59; and t(37.69) = −0.05, p = 0.96, respectively), however the average intrapair agreement did differ between the two orders of completion (Wilcoxon Rank Sum test W = 284, p = 0.023) (see Appendix II, Table C and Figure C, in Supplementary materials). There was no effect of completion order on how the overall quality of each article was graded (t(100.62) = 0.36, p = 0.723; 95%CI of difference −0.36 to 0.51).

### Usability

Time to complete the tool, and time to reach consensus, reduced as appraisers became more familiar with the tool. It took appraisers a median of 55 minutes (IQR = 43 – 59 minutes) to complete the AMSTAR-PF appraisal for the first two articles and a median of 33 minutes (IQR = 27 – 40 minutes) for the next six articles.

It took pairs a median of 18 minutes (IQR = 11-25 minutes) to reach consensus for the first two articles, and 9 minutes (IQR = 5 – 12 minutes) for the final six articles. There were no instances where pairs were unable to reach consensus. See Figure 3 and Table 3 for more details.

**Table 3.**
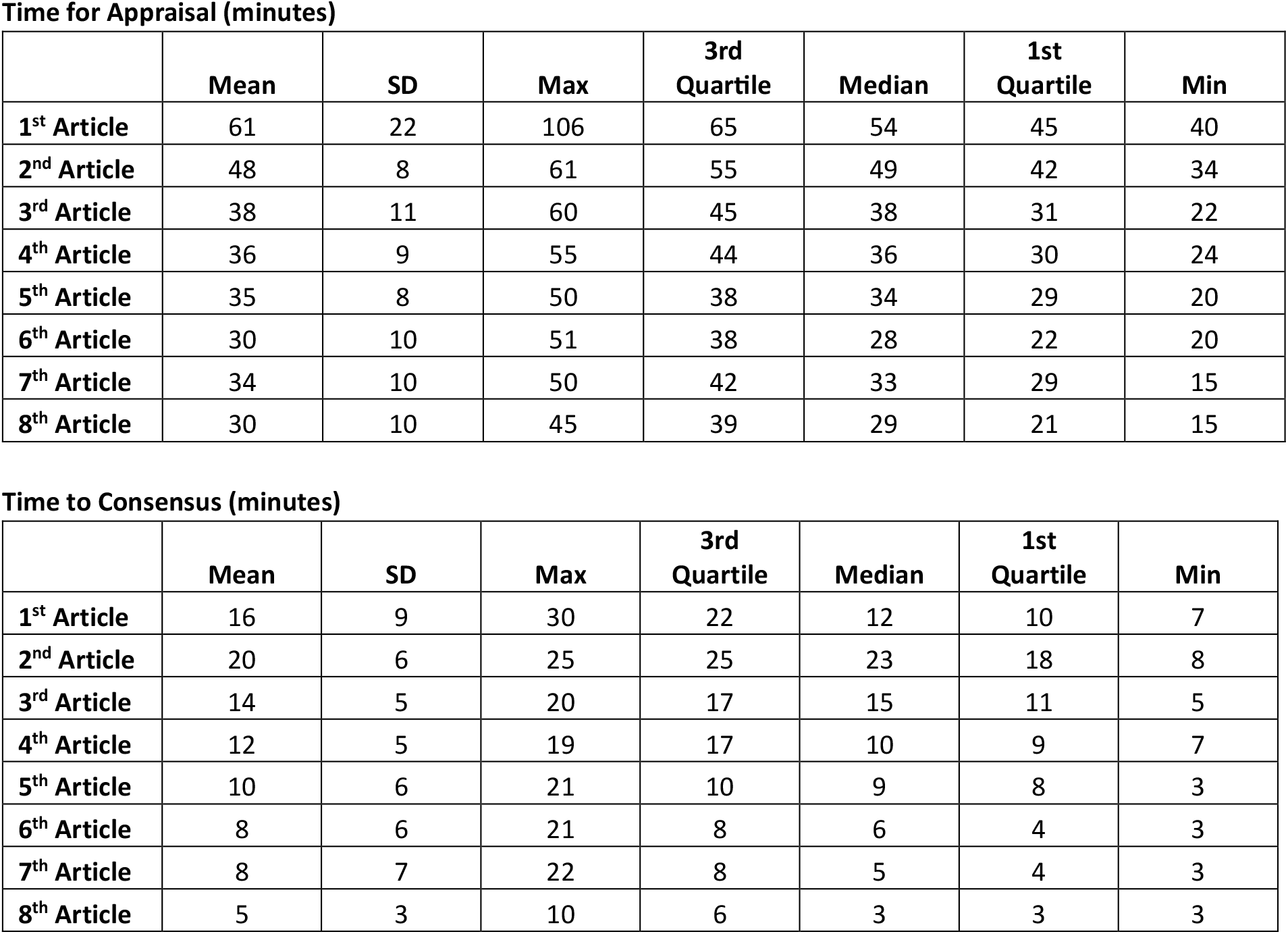
Timings for appraisal (top table) and consensus (bottom table) for each article in order of completion. All values in minutes.

**Time for Appraisal (minutes)**
|  | Mean | SD | Max | 3rd Quartile | Median | 1st Quartile | Min |
| --- | --- | --- | --- | --- | --- | --- | --- |
| **1<sup>st</sup> Article** | 61 | 22 | 106 | 65 | 54 | 45 | 40 |
| **2<sup>nd</sup> Article** | 48 | 8 | 61 | 55 | 49 | 42 | 34 |
| **3<sup>rd</sup> Article** | 38 | 11 | 60 | 45 | 38 | 31 | 22 |
| **4<sup>th</sup> Article** | 36 | 9 | 55 | 44 | 36 | 30 | 24 |
| **5<sup>th</sup> Article** | 35 | 8 | 50 | 38 | 34 | 29 | 20 |
| **6<sup>th</sup> Article** | 30 | 10 | 51 | 38 | 28 | 22 | 20 |
| **7<sup>th</sup> Article** | 34 | 10 | 50 | 42 | 33 | 29 | 15 |
| **8<sup>th</sup> Article** | 30 | 10 | 45 | 39 | 29 | 21 | 15 |

|  | Mean | SD | Max | 3rd Quartile | Median | 1st Quartile | Min |
| --- | --- | --- | --- | --- | --- | --- | --- |
| **1<sup>st</sup> Article** | 16 | 9 | 30 | 22 | 12 | 10 | 7 |
| **2<sup>nd</sup> Article** | 20 | 6 | 25 | 25 | 23 | 18 | 8 |
| **3<sup>rd</sup> Article** | 14 | 5 | 20 | 17 | 15 | 11 | 5 |
| **4<sup>th</sup> Article** | 12 | 5 | 19 | 17 | 10 | 9 | 7 |
| **5<sup>th</sup> Article** | 10 | 6 | 21 | 10 | 9 | 8 | 3 |
| **6<sup>th</sup> Article** | 8 | 6 | 21 | 8 | 6 | 4 | 3 |
| **7<sup>th</sup> Article** | 8 | 7 | 22 | 8 | 5 | 4 | 3 |
| **8<sup>th</sup> Article** | 5 | 3 | 10 | 6 | 3 | 3 | 3 |

**Figure 3.**
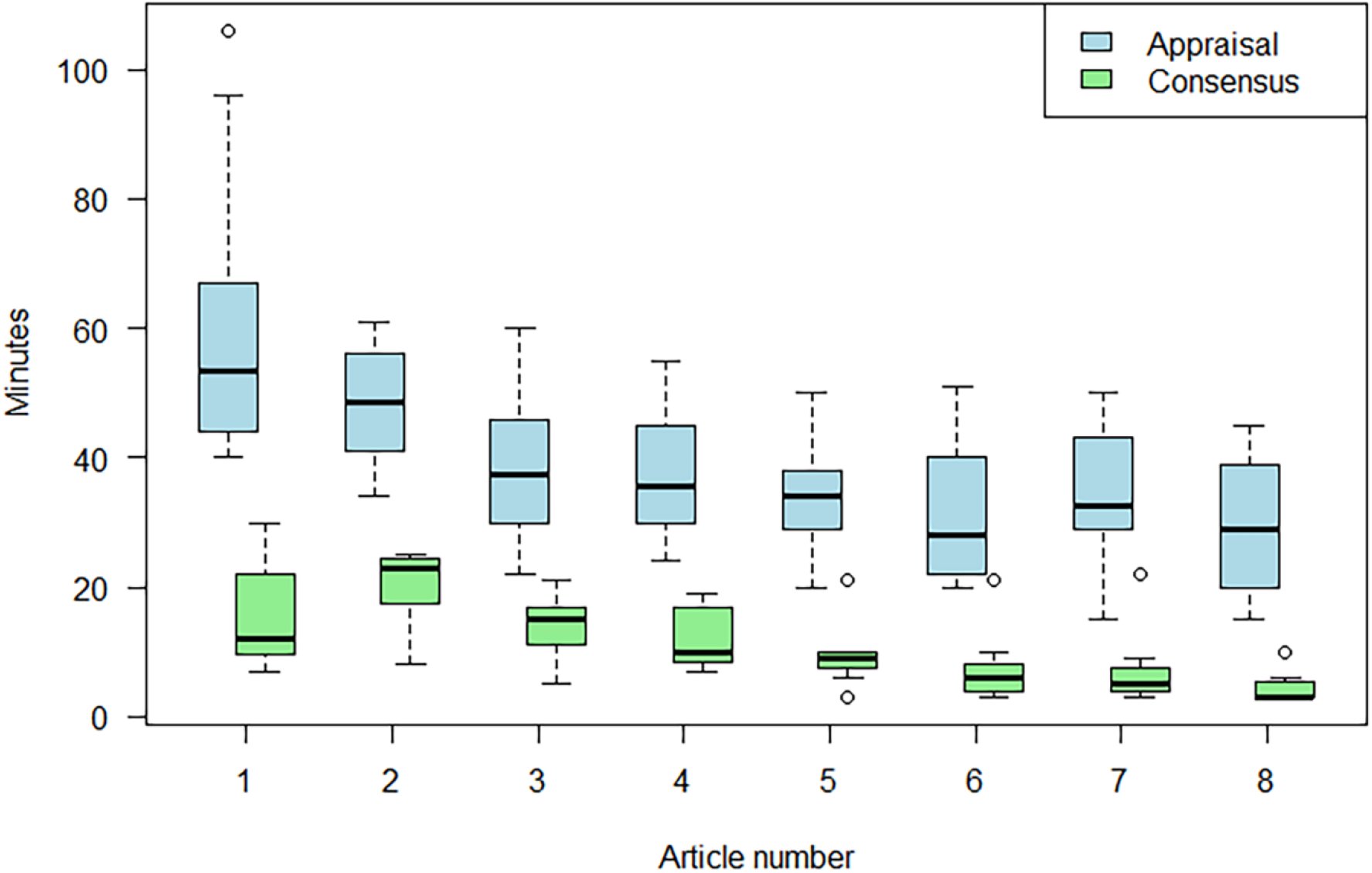
Timings to complete appraisal, and timings to complete consensus, for each article in order of completion. Box represents IQR, horizontal line is the median, and the dotted lines the range. Outliers are represented as circles.

Completion times were not different between the seven more experienced researchers (5+ years in academic research) (median completion 39 minutes (IQR = 30-51)) and the seven more novice researchers (3 or fewer years in research) (median completion 45 minutes (IQR = 30-61)) (Mann-Whitney U test: W 1593, p = 0.76, 95%CI for difference in completion times −4.0 to 5.0).

### Modification

Some questions were highlighted, either from feedback or the agreement scores, as being more difficult to answer. Some of these were more complicated, or subjective, questions, eg, *whether adequate investigation of small study effects occurred*, or *was the level of certainty around key findings described*. Other questions were more straightforward, and the poor agreement was seen as resulting from a lack of adequate guidance. Examples of this include question 2b, *Does the review justify any deviations from the protocol?* Part of the reason for this may have been subjectivity in what constituted *justification* of deviations, but partly also variation in how people responded if there was no protocol, or no publicly available protocol. Specific recommendations were added to the guidance notes to standardise this. Similarly, Q13, *Does the review report any potential sources of conflict of interest, both in the individual studies included in the review and among the review author team, including any funding received?* received low agreement scores, despite the signalling points showing high agreement. In our sample, it was common for articles to report conflicts and funding from the review authors, but not the included studies, and there was a high level of concordance with these being identified by appraisers. Some appraisers, however, routinely marked this situation as PY, others as PN, and some varied between PY and PN without any obvious rationale. More detail was included in the guidance notes to standardise responses in this situation. Similar modifications were made in other questions to try to provide further clarity or more specific guidance, however the overarching theme of all questions remained the same, and overall changes were relatively minor. Appendix III in the supplementary material compares the wording of the questions in the tested and final versions of AMSTAR-PF.

## Discussion

We aimed to determine agreement and usability of AMSTAR-PF using (i) measures of interrater, inter-pair, and intrapair agreement, and (ii) time to complete appraisal using the tool by researchers with a diverse range of research experience. Our testing of AMSTAR-PF found agreement scores that ranged from slight to almost perfect. Where agreement was lacking, changes and additions were made to the guidance notes of the tool in order to provide further clarity and improve agreement, and the wording of some questions was altered slightly in response to feedback.

The measures of agreement we calculated for AMSTAR-PF appear comparable to measures from other established tools and better than some. Direct comparison of agreeability scores is difficult, however, due to a range of measures and weightings used, and differences in testing protocols, the papers assessed, and appraisers. Published ranges for Gwet’s AC for ROBIS (Risk of Bias In Systematic reviews) (22) and AMSTAR 2 (23), for instance, range between 0.05 to 1.00 for interrater (between 2,3 or 4 raters) agreement (24-26), and −0.21 to 0.74 for inter-pair (24). AC statistics for intrapair and inter-pair agreement for the overall risk of bias using ROBINS-I (Risk Of Bias In Non-randomised Studies - of Interventions (27)) were 0.00 and 0.07, and using ROB-NRSE (Risk Of Bias for Non-Randomised Studies of Exposures) 0.11 and 0.00, respectively (28). Published benchmarked agreements when using Kappa range from no agreement to moderate agreement for a range of tools, including AMSTAR 2(25, 29, 30), ROBIS(25, 29-31), ROB 2 (Revised Cochrane Risk of Bias tool for randomised trials (32))(33, 34), ROBINS-I(35), and PROBAST (Prediction model Risk Of Bias ASsessment Tool (36, 37).

The time taken to complete an appraisal using AMSTAR-PF was also comparable to other tools. For example, documented time to complete appraisal tools include 19-35 minutes (24) for AMSTAR 2, 24-28 minutes for ROBIS (24), 28-168 minutes for ROB 2 (33, 34, 38), 27 minutes (35) and 48 minutes (28) for ROBINS-I, and 37 minutes for ROB-NRSE (28). While different articles, appraisers, and tools will contribute to variability in completion times, some of the difference may also be explained by variations in the protocol used. Some studies (eg, (24, 30)) used a protocol similar to ours, whereby only the time taken using the tool was recorded. Other studies (eg, (28, 33, 35)) included the time taken to read the review, which we did not. We decided against this because we considered it removed the impact of the length of article and appraisers’ reading speeds on our results. Our focus was on the time to use the tool, not the complete time burden of appraising articles more generally.

Our testing showed that intrapair agreeability tended to be stronger than inter-pair or interrater agreeability. This may point to a benefit in ensuring teams take time to discuss different aspects of the papers under review, and the relation to their field or review question, in order to better standardise responses and align interpretation. In our testing procedure this occurred after two papers had been appraised, but there may be benefit in also meeting prior to appraising any articles if potential issues are already known or envisioned beforehand. Guides such as the Cochrane handbook recommend planning and piloting appraisals (14).

Familiarity with the tool (and perhaps prognostic factor research and systematic reviews more generally) may reduce time to complete appraisals and gain consensus. Usability data showed improved timing as people progressed through the allocated papers, irrespective of the order of completion.

To our knowledge, AMSTAR-PF is the only tool specifically designed for reviews of prognostic factor studies, but other more general tools have been used. Most published systematic reviews undertake some form of quality assessment (39), but these vary markedly; one review found that 54 combinations of assessment tools were used across a sample of 309 reviews (39). While there are similarities in aims and constructs in many of these tools, previous research has shown that the use of different tools may lead to differing, and indeed sometimes opposite, conclusions of quality (40).

Strengths of this study include: the lodging of a full protocol prior to data collection and clearly stipulating when we deviated from that protocol; the number of appraisers involved; the diversity of experience levels; and the diversity in review topics. Articles were standardised across all appraisers to ensure a broader understanding of inter-pair agreement, and the counterbalanced order of completion helped to ensure there were direct comparisons amongst pairs who completed the same order, while also allowing comparison to detect an order effect on appraisals. Appraisers convened in the early stages of the review, which is a recommended method when applying appraisal tools in research (14).

A potential limitation to this testing protocol was the lack of prognostic factor experts in the appraiser group. Including methodological experts may have added more information and a valuable comparison to the results presented in this paper, and may be a topic for further research. We considered, however, that in practice it is often research students and early career researchers who do the bulk of the quality appraisal component of systematic reviews and umbrella reviews, albeit often with expert/s in the team they can consult when needed. Similarly, the use of articles outside the appraisers’ topic knowledge may be considered a limitation. We consider, however, that this approach has ecological validity because it is common for junior researchers to be involved in quality appraisal for reviews, which often will be on topics outside their expertise. Furthermore, we considered that if appraisers could appropriately use the tool on reviews of unfamiliar topics, they should be at least as comfortable and efficient on reviews in more familiar subject areas. All appraisers had a Cochrane review as their initial article to appraise. We acknowledge that in practice this will not always be the case, but believed that the benefit of including Cochrane and non-Cochrane reviews, and being able to compare agreement in a standardised order, outweighed this limitation. Using Landis and Koch’s benchmarks for Gwet’s AC scores is a potential limitation and has been criticised (eg, (41)), however they are commonly used to help interpret Gwet’s ACs, in part due to a lack of clear alternatives. We chose this approach to provide an interpretation of the agreement scores, but mitigated the potential limitation by using standard error to calculate cumulative probabilities, as recommended by Gwet (18), which give more conservative interpretations of the benchmarks than simply reporting where on Landis and Koch’s scale the AC score lies. We have also calculated Fleiss’ and Cohen’s Kappa and benchmarked these as a comparison (Appendix IV in the supplementary material). It is possible that including more articles for appraisal may have further improved efficiency or agreeability, but we arbitrarily chose eight as a number that would provide a good indication of change in efficiency or agreement with time, allow an assortment of different article topics and analyses, and be manageable for appraisers. Finally, we did not contact authors of the included reviews in situations where reporting was unclear or additional data may have been beneficial in coming to a judgement; meaning there were potentially more areas of uncertainty than there would be if a complete umbrella review was being performed, as opposed to just quality appraisal. That is, adding this step in practice may improve agreeability and thus performance of the AMSTAR-PF tool.

## Conclusion

AMSTAR-PF displays an acceptable level of usability and agreement, offering a valuable tool for those appraising the quality of systematic reviews of prognostic factors. There may be benefit for appraiser pairs to meet prior to commencing quality appraisal, to ensure standardised interpretation of the tool and how the different domains may be relevant to the topic area of the reviews under appraisal.

## Data Availability

All data produced in the present study are available upon reasonable request to the authors

## Statement of contribution

Contributors: MLH, NEO’C, RR, KGM, BJS, LH, JAAD, NS, SBW, and GLM developed and/or refined the tool. RPA, CB, SMC, GAF, ARG, KMH, MLH, AMH, ELK, IK, HBL, EM, BM, KP, DAV, SBW, EW, LKW, EMW, MKW, MVW were involved in testing different iterations of the tool. MLH analysed the data and wrote the first draft of the manuscript. All authors critically revised the manuscript and approved the final version.

MLH is the guarantor of the article. The corresponding author attests that all listed authors meet authorship criteria and that no others meeting the criteria have been omitted.

## Data Availability

The data that support the findings of this study are available from the corresponding author, NEO’C, upon reasonable request.

## References

1. Riley RD, van der Windt DA, Croft P, Moons KGM. Prognosis Research in Healthcare: Concepts, Methods, and Impact: Oxford University Press; 2019. 376 p.

2. Hoffmann F, Allers K, Rombey T, Helbach J, Hoffmann A, Mathes T, et al. Nearly 80 systematic reviews were published each day: Observational study on trends in epidemiology and reporting over the years 2000-2019. Journal of Clinical Epidemiology. 2021;138:1–11.

3. Riley RD, Hayden JA, Steyerberg EW, Moons KG, Abrams K, Kyzas PA, et al. Prognosis Research Strategy (PROGRESS) 2: prognostic factor research. PLoS Med. 2013;10(2):e1001380.

4. Kent P, Cancelliere C, Boyle E, Cassidy JD, Kongsted A. A conceptual framework for prognostic research. BMC Med Res Methodol. 2020;20(1):172.

5. Henry ML, O’Connell NE, Riley RD, Moons KG, Shea BJ, Hooft L, et al. AMSTAR-PF: a critical appraisal tool for systematic reviews of prognostic factor studies. medRxiv [Preprint], 2025.

6. Hayden JA, Wilson MN, Riley RD, Iles R, Pincus T, Ogilvie R. Individual recovery expectations and prognosis of outcomes in non-specific low back pain: prognostic factor review. Cochrane Database Syst Rev. 2019;2019(11).

7. Pinheiro MB, Ferreira ML, Refshauge K, Maher CG, Ordoñana JR, Andrade TB, et al. Symptoms of depression as a prognostic factor for low back pain: a systematic review. The Spine Journal. 2016;16(1):105–16.

8. Wijnands AM, de Jong ME, Lutgens M, Hoentjen F, Elias SG, Oldenburg B, et al. Prognostic Factors for Advanced Colorectal Neoplasia in Inflammatory Bowel Disease: Systematic Review and Meta-analysis. Gastroenterology. 2021;160(5):1584–98.

9. Aldin A, Umlauff L, Estcourt LJ, Collins G, Moons KG, Engert A, et al. Interim PET-results for prognosis in adults with Hodgkin lymphoma: a systematic review and meta-analysis of prognostic factor studies. Cochrane Database Syst Rev. 2020;1(1):CD012643.

10. Puig J, Ellis MJ, Kornelsen J, Figley TD, Figley CR, Daunis IEP, et al. Magnetic Resonance Imaging Biomarkers of Brain Connectivity in Predicting Outcome after Mild Traumatic Brain Injury: A Systematic Review. J Neurotrauma. 2020;37(16):1761–76.

11. Mercier E, Tardif P-A, Cameron PA, Batomen Kuimi BL, Emond M, Moore L, et al. Prognostic value of S-100β protein for prediction of post-concussion symptoms after a mild traumatic brain injury: systematic review and meta-analysis. Journal of Neurotrauma. 2018;35(4):609–22.

12. Izcovich A, Ragusa MA, Tortosa F, Lavena Marzio MA, Agnoletti C, Bengolea A, et al. Prognostic factors for severity and mortality in patients infected with COVID-19: A systematic review. PloS one. 2020;15(11):e0241955.

13. Maglietta G, Diodati F, Puntoni M, Lazzarelli S, Marcomini B, Patrizi L, et al. Prognostic factors for post-COVID-19 syndrome: a systematic review and meta-analysis. Journal of clinical medicine. 2022;11(6):1541.

14. Boutron I PM, Higgins JPT, Altman DG, Lundh A, Hróbjartsson A. Chapter 7: Considering bias and conflicts of interest among the included studies [last updated August 2022]. In: Higgins JPT TJ, Chandler J, Cumpston M, Li T, Page MJ, Welch VA, editor. Cochrane Handbook for Systematic Reviews of Interventions version 65: Cochrane; 2024.

15. Gwet KL. Computing inter-rater reliability and its variance in the presence of high agreement. Br J Math Stat Psychol. 2008;61(Pt 1):29–48.

16. Cicchetti DV, Feinstein AR. High agreement but low kappa: II. Resolving the paradoxes. J Clin Epidemiol. 1990;43(6):551–8.

17. Feinstein AR, Cicchetti DV. High agreement but low kappa: I. The problems of two paradoxes. J Clin Epidemiol. 1990;43(6):543–9.

18. Gwet KL. Handbook of Inter-Rater Reliability: The Definitive Guide to Measuring the Extent of Agreement Among Raters, 4th Edition 4ed: Advanced Analytics, LLC; 2014. 428 p.

19. Landis JR, Koch GG. The measurement of observer agreement for categorical data. Biometrics. 1977;33(1):159–74.

20. Petticrew M, Wilson P, Wright K, Song F. Quality of Cochrane reviews. Quality of Cochrane reviews is better than that of non-Cochrane reviews. Bmj. 2002;324(7336):545.

21. Moseley AM, Elkins MR, Herbert RD, Maher CG, Sherrington C. Cochrane reviews used more rigorous methods than non-Cochrane reviews: survey of systematic reviews in physiotherapy. Journal of Clinical Epidemiology. 2009;62(10):1021–30.

22. Whiting P, Savovic J, Higgins JP, Caldwell DM, Reeves BC, Shea B, et al. ROBIS: A new tool to assess risk of bias in systematic reviews was developed. J Clin Epidemiol. 2016;69:225–34.

23. Shea BJ, Reeves BC, Wells G, Thuku M, Hamel C, Moran J, et al. AMSTAR 2: a critical appraisal tool for systematic reviews that include randomised or non-randomised studies of healthcare interventions, or both. BMJ. 2017;358:j4008.

24. Gates M, Gates A, Duarte G, Cary M, Becker M, Prediger B, et al. Quality and risk of bias appraisals of systematic reviews are inconsistent across reviewers and centers. J Clin Epidemiol. 2020;125:9–15.

25. Lorenz RC, Matthias K, Pieper D, Wegewitz U, Morche J, Nocon M, et al. A psychometric study found AMSTAR 2 to be a valid and moderately reliable appraisal tool. Journal of Clinical Epidemiology. 2019;114:133–40.

26. Perry R, Whitmarsh A, Leach V, Davies P. A comparison of two assessment tools used in overviews of systematic reviews: ROBIS versus AMSTAR-2. Systematic Reviews. 2021;10(1):273.

27. Sterne JA, Hernán MA, Reeves BC, Savović J, Berkman ND, Viswanathan M, et al. ROBINS-I: a tool for assessing risk of bias in non-randomised studies of interventions. BMJ. 2016;355:i4919.

28. Jeyaraman MM, Rabbani R, Copstein L, Robson RC, Al-Yousif N, Pollock M, et al. Methodologically rigorous risk of bias tools for nonrandomized studies had low reliability and high evaluator burden. Journal of Clinical Epidemiology. 2020;128:140–7.

29. Pieper D, Puljak L, González-Lorenzo M, Minozzi S. Minor differences were found between AMSTAR 2 and ROBIS in the assessment of systematic reviews including both randomized and nonrandomized studies. Journal of Clinical Epidemiology. 2019;108:26–33.

30. Lee SWH. What tool do undergraduate pharmacy students prefer when grading systematic review evidence: AMSTAR-2 or ROBIS? Cochrane Evidence Synthesis and Methods. 2023;1(6).

31. Bühn S, Mathes T, Prengel P, Wegewitz U, Ostermann T, Robens S, et al. The risk of bias in systematic reviews tool showed fair reliability and good construct validity. Journal of Clinical Epidemiology. 2017;91:121–8.

32. Sterne JAC, Savović J, Page MJ, Elbers RG, Blencowe NS, Boutron I, et al. RoB 2: a revised tool for assessing risk of bias in randomised trials. BMJ. 2019;366:4898.

33. Minozzi S, Cinquini M, Gianola S, Gonzalez-Lorenzo M, Banzi R. The revised Cochrane risk of bias tool for randomized trials (RoB 2) showed low interrater reliability and challenges in its application. Journal of Clinical Epidemiology. 2020;126:37–44.

34. Minozzi S, Dwan K, Borrelli F, Filippini G. Reliability of the revised Cochrane risk-of-bias tool for randomised trials (RoB2) improved with the use of implementation instruction. Journal of Clinical Epidemiology. 2022;141:99–105.

35. Minozzi S, Cinquini M, Gianola S, Castellini G, Gerardi C, Banzi R. Risk of bias in nonrandomized studies of interventions showed low inter-rater reliability and challenges in its application. Journal of Clinical Epidemiology. 2019;112:28–35.

36. Wolff RF, Moons KGM, Riley RD, Whiting PF, Westwood M, Collins GS, et al. PROBAST: A Tool to Assess the Risk of Bias and Applicability of Prediction Model Studies. Ann Intern Med. 2019;170(1):51–8.

37. Langenhuijsen LFS, Janse RJ, Venema E, Kent DM, van Diepen M, Dekker FW, et al. Systematic metareview of prediction studies demonstrates stable trends in bias and low PROBAST inter-rater agreement. Journal of Clinical Epidemiology. 2023;159:159–73.

38. Crocker TF, Lam N, Jordão M, Brundle C, Prescott M, Forster A, et al. Risk-of-bias assessment using Cochrane’s revised tool for randomized trials (RoB 2) was useful but challenging and resource-intensive: observations from a systematic review. Journal of Clinical Epidemiology. 2023;161:39–45.

39. Seehra J, Pandis N, Koletsi D, Fleming PS. Use of quality assessment tools in systematic reviews was varied and inconsistent. Journal of Clinical Epidemiology. 2016;69:179-84.e5.

40. Losilla J-M, Oliveras I, Marin-Garcia JA, Vives J. Three risk of bias tools lead to opposite conclusions in observational research synthesis. Journal of Clinical Epidemiology. 2018;101:61–72.

41. Vach W, Gerke O. Gwet’s AC1 is not a substitute for Cohen’s kappa - A comparison of basic properties. MethodsX. 2023;10:102212.

