## Supplementary Files for "Agreeability testing of AMSTAR-PF, a tool for quality appraisal of systematic reviews of prognostic factor studies"

### Supplementary Material

#### Appendix I – AMSTAR-PF

| | |
|---|---|
| **1** | Did the review clearly define the research question, including the relevant components of PICOTS? |
| **2a** | Did the review state that a protocol was registered prior to the conduct of the review, and is that registration publicly available? |
| **2b** | Did the review justify any deviations from the protocol? |
| **3** | Did the review report the types of prognostic factor studies eligible for inclusion in the review, eg, which primary study designs were eligible? |
| **4** | Did the review use a comprehensive search strategy to identify relevant studies? |
| **5** | Was a rigorous process followed when evaluating studies for inclusion into the review? |
| **6** | Did the review list all excluded studies that were included in the full text screening, and the reasons for those exclusions? |
| **7a** | Was a rigorous process followed when performing data extraction for each included study? |
| **7b** | Did the review describe the included studies in adequate detail? |
| **7c** | Did the review use appropriate techniques for calculating or obtaining the Prognostic Factor effect estimates and their precision, in situations they were not fully reported in the primary studies? |
| **8a** | Was a rigorous process followed when performing risk of bias (RoB) assessment for each included study? |
| **8b** | Did the review use an appropriate technique for assessing the risk of bias (RoB) of individual studies that were included in the review? |
| **9a** | If synthesis was performed, did the approach taken ensure the interpretability of results? |
| **9b** | If meta-analysis was performed, did the review use appropriate analysis methods? |
| **10** | If the review performed quantitative data synthesis, did it also carry out an adequate investigation of small study effects? |
| **11** | Did the review account for risk of bias (RoB) in individual studies when interpreting/discussing the results of the review? |
| **12** | Did the review discuss any heterogeneity observed in its results? |
| **13** | Did the review report any potential sources of conflict of interest, both in the individual studies included in the review and among the review author team, including any funding received? |
| **14** | Did the review address the level of certainty around their key findings and use appropriate methods to come to an overall certainty judgement each outcome? |

**Summary of AMSTAR-PF questions** (from Henry ML, O'Connell NE, Riley RD, Moons KG, Shea BJ, Hooft L, et al. AMSTAR-PF: a critical appraisal tool for systematic reviews of prognostic factor studies. *medRxiv* [Preprint], 2025). Note that the testing used a slightly different version of the tool; see Appendix III in these supplementary files for a comparison of the AMSTAR-PF version used for testing compared to the published version shown here.

### Appendix II – Subgroup comparisons of article agreement

| | First two articles | | Last six articles | | Last two articles | |
|---|---|---|---|---|---|---|
| Question | AC | SEM | AC | SEM | AC | SEM |
| **1** | 1.00 | 0.00 | 0.80 | 0.14 | 0.85 | 0.19 |
| **2a** | 0.83 | 0.23 | 0.92 | 0.07 | 0.76 | 0.17 |
| **2b** | 0.49 | 0.36 | 0.55 | 0.25 | 0.67 | 0.36 |
| **3** | 0.97 | 0.04 | 0.78 | 0.15 | 0.83 | 0.21 |
| **4** | 0.74 | 0.34 | 0.83 | 0.12 | 0.93 | 0.09 |
| **5** | 0.87 | 0.17 | 0.82 | 0.14 | 0.87 | 0.20 |
| **6** | 0.69 | 0.26 | 0.88 | 0.07 | 0.92 | 0.11 |
| **7a** | 0.87 | 0.17 | 0.94 | 0.04 | 0.97 | 0.04 |
| **7b** | 0.84 | 0.21 | 0.81 | 0.13 | 0.82 | 0.24 |
| **7c** | 0.42 | 0.25 | 0.47 | 0.26 | 0.67 | 0.00 |
| **8a** | 0.78 | 0.13 | 0.92 | 0.06 | 0.97 | 0.04 |
| **8b** | 0.93 | 0.09 | 0.95 | 0.05 | 1.00 | 0.00 |
| **9a** | 0.84 | 0.17 | 0.54 | 0.21 | 0.67 | 0.17 |
| **9b** | 0.52 | 0.34 | 0.80 | 0.18 | 0.76 | 0.25 |
| **10** | 0.75 | 0.09 | 0.54 | 0.21 | 0.75 | 0.27 |
| **11** | 0.87 | 0.17 | 0.61 | 0.22 | 0.85 | 0.19 |
| **12** | 0.67 | 0.18 | 0.59 | 0.25 | 0.68 | 0.34 |
| **13** | 0.74 | 0.26 | 0.81 | 0.12 | 0.95 | 0.06 |
| **14** | 0.81 | 0.26 | 0.52 | 0.24 | 0.77 | 0.30 |
| **Final** | 0.63 | 0.19 | 0.70 | 0.14 | 0.87 | 0.17 |
| **Mean** | **0.76** | | **0.74** | | **0.83** | |
| **SEM** | 0.03 | | 0.04 | | 0.02 | |
| **SD** | 0.16 | | 0.16 | | 0.11 | |
| **min** | 0.42 | | 0.47 | | 0.67 | |
| **max** | 1.00 | | 0.95 | | 1.00 | |
| **Benchmark** | 0.6-0.8 | | 0.6-0.8 | | 0.6-0.8 | |

**Table A: Average Gwet's AC for the intrapair agreement for the first two, final six, and final two articles appraised**. Benchmark interpretation is calculated using 95% cumulative probabilities for Landis and Koch's benchmark categories, < 0, Poor; 0.0-0.2, Slight; 0.2-0.4, Fair; 0.4-0.6, Moderate; 0.6-0.8, Substantial; and 0.8-1.0, Almost Perfect.

AC, Gwet's Agreement Coefficient; min, minimum; max, maximum; SEM, Standard Error of the Mean; SD, Standard Deviation

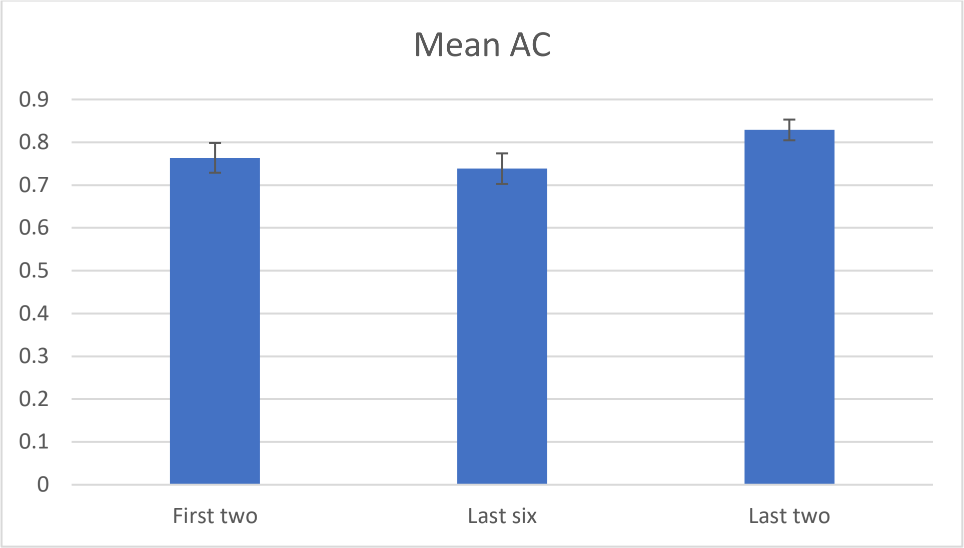

**Figure A: Average intrapair Gwet's AC for the first two, last six, and last two articles appraised.** Error bars are standard error of the mean.

| | Interrater | | | | Inter-pair | | | | Intrapair | | | |
|---|---|---|---|---|---|---|---|---|---|---|---|---|
| | Cochrane | | Non-Cochrane | | Cochrane | | Non-Cochrane | | Cochrane | | Non-Cochrane | |
| Question | AC | SE | AC | SE | AC | SE | AC | SE | Avg AC | SEM | Avg AC | SEM |
| 1 | 0.97 | 0.03 | 0.57 | 0.11 | 0.95 | 0.06 | 0.57 | 0.10 | 0.97 | 0.03 | 0.79 | 0.07 |
| 2a | 0.39 | 0.27 | 0.99 | 0.01 | 0.20 | 0.17 | 1.00 | 0.00 | 0.67 | 0.14 | 0.99 | 0.01 |
| 2b | 0.85 | 0.00 | 0.13 | 0.04 | 0.70 | 0.00 | 0.17 | 0.05 | 0.84 | 0.10 | 0.45 | 0.10 |
| 3 | 1.00 | 0.00 | 0.70 | 0.14 | 1.00 | 0.00 | 0.83 | 0.12 | 1.00 | 0.00 | 0.77 | 0.10 |
| 4 | 0.87 | 0.08 | 0.67 | 0.04 | 0.77 | 0.00 | 0.76 | 0.06 | 0.90 | 0.05 | 0.76 | 0.08 |
| 5 | 0.92 | 0.08 | 0.79 | 0.10 | 0.90 | 0.12 | 0.81 | 0.13 | 0.90 | 0.07 | 0.81 | 0.07 |
| 6 | 0.81 | 0.15 | 0.81 | 0.05 | 0.76 | 0.14 | 0.91 | 0.02 | 0.89 | 0.05 | 0.86 | 0.06 |
| 7a | 0.91 | 0.11 | 0.79 | 0.07 | 0.90 | 0.12 | 0.80 | 0.08 | 0.93 | 0.04 | 0.91 | 0.04 |
| 7b | 0.97 | 0.03 | 0.62 | 0.09 | 0.95 | 0.06 | 0.66 | 0.05 | 0.97 | 0.03 | 0.76 | 0.08 |
| 7c | 0.45 | 0.16 | 0.14 | 0.06 | 0.47 | 0.24 | 0.17 | 0.05 | 0.67 | 0.17 | 0.38 | 0.07 |
| 8a | 0.86 | 0.10 | 0.78 | 0.12 | 0.87 | 0.16 | 0.81 | 0.12 | 0.93 | 0.04 | 0.88 | 0.06 |
| 8b | 1.00 | 0.00 | 0.86 | 0.11 | 1.00 | 0.00 | 0.87 | 0.12 | 1.00 | 0.00 | 0.93 | 0.03 |
| 9a | 0.81 | 0.20 | 0.28 | 0.11 | 0.75 | 0.27 | 0.23 | 0.10 | 0.92 | 0.08 | 0.51 | 0.13 |
| 9b | 0.85 | 0.00 | 0.51 | 0.12 | 1.00 | 0.00 | 0.60 | 0.09 | 0.84 | 0.10 | 0.69 | 0.07 |
| 10 | 0.55 | 0.32 | 0.16 | 0.11 | 0.60 | 0.44 | 0.15 | 0.11 | 0.76 | 0.11 | 0.54 | 0.11 |
| 11 | 0.95 | 0.00 | 0.09 | 0.10 | 1.00 | 0.00 | 0.13 | 0.06 | 0.93 | 0.04 | 0.59 | 0.13 |
| 12 | 0.52 | 0.08 | 0.24 | 0.12 | 0.47 | 0.00 | 0.33 | 0.11 | 0.68 | 0.11 | 0.57 | 0.09 |
| 13 | 0.97 | 0.03 | 0.50 | 0.10 | 1.00 | 0.00 | 0.39 | 0.07 | 0.97 | 0.03 | 0.78 | 0.06 |
| 14 | 1.00 | 0.00 | 0.10 | 0.09 | 1.00 | 0.00 | 0.14 | 0.14 | 1.00 | 0.00 | 0.45 | 0.10 |
| Final | 0.89 | 0.00 | 0.44 | 0.06 | 1.00 | 0.00 | 0.38 | 0.06 | 0.85 | 0.08 | 0.70 | 0.09 |
| Mean | **0.83** | | **0.51** | | **0.81** | | **0.54** | | **0.88** | | **0.71** | |
| SEM | 0.04 | | 0.07 | | 0.05 | | 0.07 | | 0.02 | | 0.04 | |
| SD | 0.19 | | 0.29 | | 0.23 | | 0.30 | | 0.11 | | 0.18 | |
| min | 0.39 | | 0.09 | | 0.20 | | 0.13 | | 0.67 | | 0.38 | |
| max | 1.00 | | 0.99 | | 1.00 | | 1.00 | | 1.00 | | 0.99 | |
| B-M | 0.6-0.8 | | 0.4-0.6 | | 0.6-0.8 | | 0.4-0.6 | | 0.8-1.0 | | 0.6-0.8 | |

**Table B: Gwet's AC for Interrater, Inter-pair, and Intrapair agreement for the Cochrane and non-Cochrane articles.** Benchmark interpretation is calculated using 95% cumulative probabilities for Landis and Koch's benchmark categories, < 0, Poor; 0.0-0.2, Slight; 0.2-0.4, Fair; 0.4-0.6, Moderate; 0.6-0.8, Substantial; and 0.8-1.0, Almost Perfect.

AC, Gwet's Agreement Coefficient; Avg, Average; B-M, Benchmark interpretation; min, minimum; max, maximum; SD, Standard Deviation; SE, Standard Error; SEM, Standard Error of the Mean.

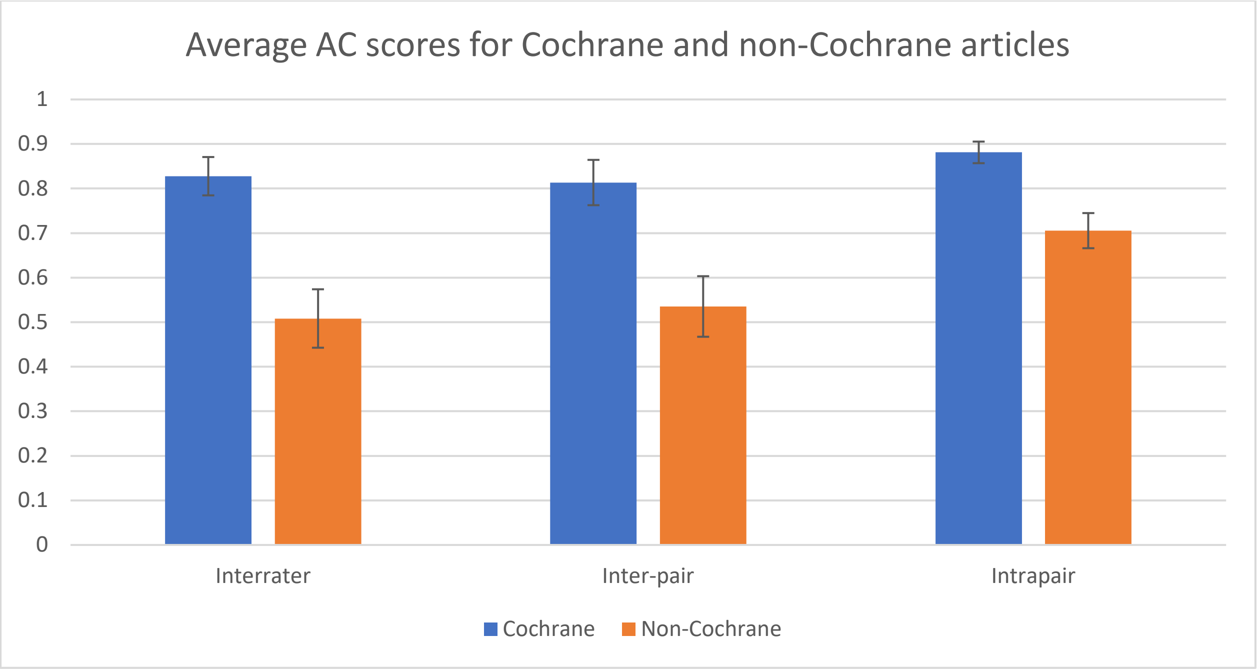

**Figure B: Average intrapair Gwet's AC for the Cochrane and non-Cochrane articles**. Error bars are standard error of the mean.

| | Interrater | | | | Inter-pair | | | | Intrapair | | | |
|---|---|---|---|---|---|---|---|---|---|---|---|---|
| | Stream 1 | | Stream 2 | | Stream 1 | | Stream 2 | | Stream 1 | | Stream 2 | |
| **Question** | **AC** | **SE** | **AC** | **SE** | **AC** | **SE** | **AC** | **SE** | **Avg AC** | **SEM** | **Avg AC** | **SEM** |
| **1** | 0.68 | 0.08 | 0.75 | 0.12 | 0.71 | 0.07 | 0.71 | 0.13 | 0.83 | 0.06 | 0.84 | 0.09 |
| **2a** | 0.88 | 0.11 | 0.85 | 0.10 | 0.84 | 0.13 | 0.83 | 0.14 | 0.98 | 0.02 | 0.84 | 0.07 |
| **2b** | 0.32 | 0.12 | 0.25 | 0.15 | 0.23 | 0.12 | 0.25 | 0.12 | 0.55 | 0.09 | 0.51 | 0.11 |
| **3** | 0.85 | 0.06 | 0.77 | 0.14 | 0.88 | 0.08 | 0.93 | 0.08 | 0.95 | 0.05 | 0.75 | 0.09 |
| **4** | 0.77 | 0.07 | 0.65 | 0.09 | 0.83 | 0.07 | 0.69 | 0.02 | 0.86 | 0.04 | 0.75 | 0.07 |
| **5** | 0.87 | 0.07 | 0.80 | 0.10 | 0.88 | 0.08 | 0.79 | 0.12 | 0.88 | 0.06 | 0.80 | 0.07 |
| **6** | 0.69 | 0.12 | 0.78 | 0.05 | 0.64 | 0.14 | 0.94 | 0.05 | 0.88 | 0.07 | 0.79 | 0.11 |
| **7a** | 0.85 | 0.06 | 0.80 | 0.08 | 0.84 | 0.08 | 0.81 | 0.10 | 0.95 | 0.01 | 0.89 | 0.06 |
| **7b** | 0.77 | 0.09 | 0.63 | 0.11 | 0.75 | 0.07 | 0.59 | 0.09 | 0.90 | 0.02 | 0.73 | 0.07 |
| **7c** | 0.12 | 0.04 | 0.30 | 0.12 | 0.09 | 0.07 | 0.37 | 0.12 | 0.51 | 0.13 | 0.40 | 0.14 |
| **8a** | 0.80 | 0.07 | 0.80 | 0.11 | 0.79 | 0.08 | 0.83 | 0.12 | 0.93 | 0.07 | 0.87 | 0.05 |
| **8b** | 0.91 | 0.07 | 0.89 | 0.09 | 0.92 | 0.08 | 0.93 | 0.08 | 0.98 | 0.02 | 0.93 | 0.03 |
| **9a** | 0.48 | 0.11 | 0.43 | 0.13 | 0.41 | 0.12 | 0.29 | 0.13 | 0.70 | 0.09 | 0.55 | 0.19 |
| **9b** | 0.68 | 0.10 | 0.64 | 0.13 | 0.80 | 0.13 | 0.66 | 0.14 | 0.70 | 0.09 | 0.75 | 0.09 |
| **10** | 0.24 | 0.14 | 0.28 | 0.10 | 0.28 | 0.17 | 0.28 | 0.14 | 0.55 | 0.15 | 0.62 | 0.05 |
| **11** | 0.43 | 0.13 | 0.42 | 0.17 | 0.36 | 0.19 | 0.42 | 0.21 | 0.89 | 0.03 | 0.54 | 0.11 |
| **12** | 0.49 | 0.14 | 0.44 | 0.14 | 0.41 | 0.14 | 0.48 | 0.14 | 0.71 | 0.00 | 0.52 | 0.08 |
| **13** | 0.70 | 0.12 | 0.50 | 0.13 | 0.47 | 0.19 | 0.55 | 0.15 | 0.87 | 0.07 | 0.73 | 0.07 |
| **14** | 0.44 | 0.19 | 0.37 | 0.23 | 0.43 | 0.19 | 0.42 | 0.25 | 0.61 | 0.15 | 0.58 | 0.08 |
| **Final** | 0.58 | 0.09 | 0.45 | 0.08 | 0.57 | 0.12 | 0.44 | 0.14 | 0.89 | 0.06 | 0.52 | 0.10 |
| **Mean** | **0.63** | | **0.59** | | **0.61** | | **0.61** | | **0.81** | | **0.70** | |
| **SEM** | 0.05 | | 0.05 | | 0.06 | | 0.05 | | 0.03 | | 0.03 | |
| **SD** | 0.23 | | 0.21 | | 0.25 | | 0.23 | | 0.15 | | 0.15 | |
| **min** | 0.12 | | 0.25 | | 0.09 | | 0.25 | | 0.51 | | 0.40 | |
| **max** | 0.91 | | 0.89 | | 0.92 | | 0.94 | | 0.98 | | 0.93 | |
| **B-M** | 0.4-0.6 | | 0.4-0.6 | | 0.4-0.6 | | 0.4-0.6 | | 0.6-0.8 | | 0.6-0.8 | |

**Table C: Gwet's AC for Interrater, Inter-pair, and Intrapair agreement for the two different orders of completion.** Benchmark interpretation is calculated using 95% cumulative probabilities for Landis and Koch's benchmark categories, < 0, Poor; 0.0-0.2, Slight; 0.2-0.4, Fair; 0.4-0.6, Moderate; 0.6-0.8, Substantial; and 0.8-1.0, Almost Perfect.

AC, Gwet's Agreement Coefficient; Avg, Average; B-M, Benchmark; min, minimum; max, maximum; SD, Standard Deviation; SE, Standard Error; SEM, Standard Error of the Mean

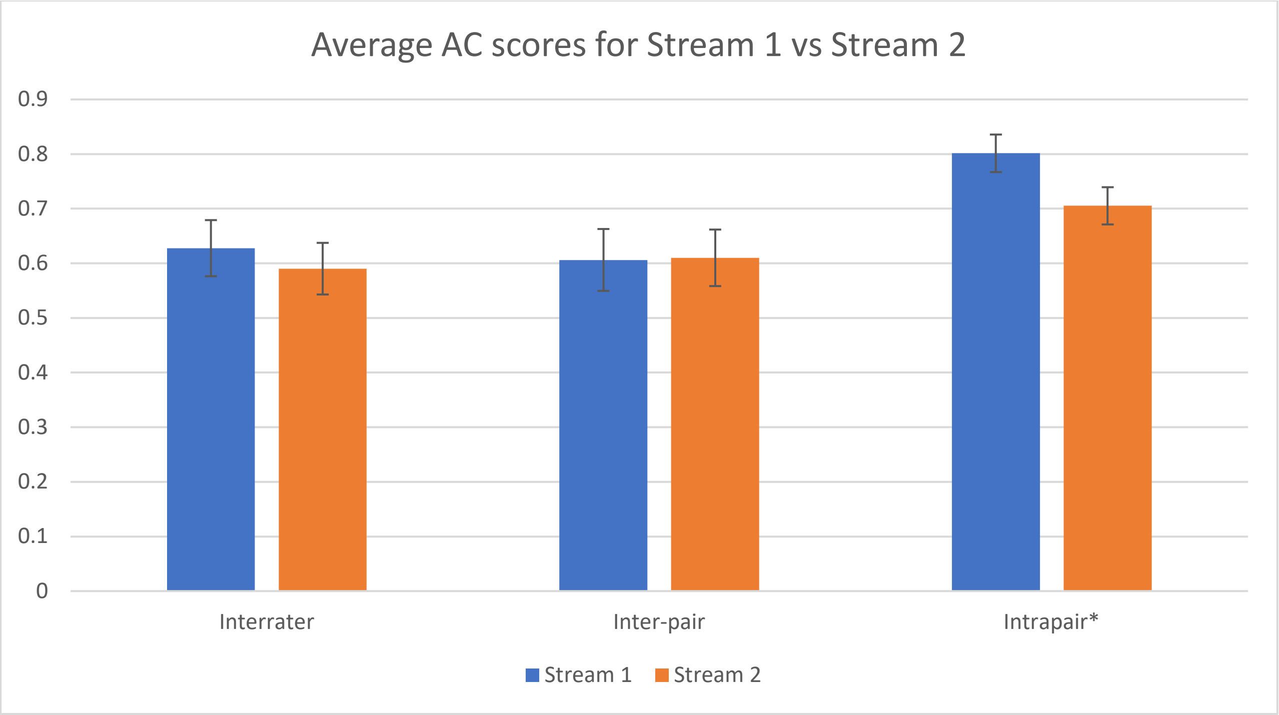

**Figure C: Average interrater, inter-pair, and intrapair Gwet's AC for two orders of completion**. Error bars are standard error of the mean. *Signifies significant difference at $p < 0.005$

**Appendix III – AMSTAR-PF – Tested version compared to the final version**

| | |
|---|---|
| **1** | ~~Does~~ Did the review clearly define the research question, including the relevant components of PICOTS? |
| **2a** | ~~Does~~ Did the review ~~contain a clear statement~~ state that a protocol was registered prior to the conduct of the review, and is that ~~the~~ registration ~~is~~ publicly available? |
| **2b** | ~~Does~~ Did the review justify any deviations from the protocol? |
| **3** | ~~Does~~ Did the review report the types of prognostic factor studies eligible for inclusion in the review, eg, which primary study designs were eligible? |
| **4** | ~~Does~~ Did the review use a comprehensive search strategy to identify relevant studies? |
| **5** | Was a rigorous process followed when evaluating studies for inclusion into the review? |
| **6** | ~~Does~~ Did the review list all excluded studies that were included in the full text screening, and the reasons for those exclusions? |
| **7a** | Was a rigorous process followed when performing data extraction for each included study? |
| **7b** | ~~Does~~ Did the review describe the included studies in adequate detail? |
| **7c** | ~~Does~~ Did the review use appropriate techniques for calculating or obtaining the Prognostic Factor effect estimates and their precision, in situations they were not fully reported in the primary studies? |
| **8a** | Was a rigorous process followed when performing risk of bias (RoB) assessment for each included study? |
| **8b** | ~~Does~~ Did the review use an appropriate technique for assessing the risk of bias (RoB) of individual studies that were included in the review? |
| **9a** | If ~~data~~ synthesis was performed, ~~does~~ did the approach taken ensure the interpretability of results? |
| **9b** | If meta-analysis was performed, ~~does~~ did the review use appropriate analysis methods? |
| **10** | If the review performed quantitative data synthesis, ~~does~~ did it also carry out an adequate investigation of small study effects? |
| **11** | ~~Does~~ Did the review account for risk of bias (RoB) in individual studies when interpreting/discussing the results of the review? |
| **12** | ~~Does~~ Did the review discuss any heterogeneity observed in its results? |
| **13** | ~~Does~~ Did the review report any potential sources of conflict of interest, both in the individual studies included in the review and among the review author team, including any funding received? |
| **14** | ~~Does~~ Did the review address the level of certainty around their key findings and use appropriate methods to come to an overall certainty judgement each outcome? |

**Summary of AMSTAR-PF questions in the final version of the tool, compared with the version tested.** Words in blue font have been added for the final version; words with strikethrough have been removed.

### Appendix IV – Kappa scores for agreement

| Question | | Interrater | | | | Inter-pair | | | | Intrapair | | | |
|---|---|---|---|---|---|---|---|---|---|---|---|---|---|
| | | F Kap | 95% CI | | I | F Kap | 95% CI | | I | C Kap | 95% CI | | I |
| 1 | Research Question | 0.27 | 0.06 | 0.49 | | 0.21 | -0.04 | 0.46 | | 0.64 | 0.42 | 0.85 | |
| 2a | Protocol Registration | 0.74 | 0.29 | 1.00 | | 0.69 | 0.19 | 1.00 | | 0.83 | 0.67 | 0.98 | |
| ^2b | Deviations from protocol | 0.27 | 0.01 | 0.53 | | 0.25 | 0.04 | 0.47 | | 0.44 | 0.24 | 0.63 | |
| 3 | Included study designs | 0.12 | 0.00 | 0.25 | | 0.27 | 0.05 | 0.49 | | 0.42 | -0.11 | 0.94 | |
| 4 | Search strategy | 0.23 | 0.04 | 0.42 | | 0.14 | -0.05 | 0.32 | | 0.40 | 0.11 | 0.68 | |
| 5 | Inclusion process | 0.20 | -0.02 | 0.41 | | 0.35 | 0.12 | 0.58 | | 0.40 | 0.06 | 0.74 | |
| 6 | Excluded studies | 0.65 | 0.37 | 0.94 | | 0.72 | 0.53 | 0.92 | | 0.76 | 0.57 | 0.95 | |
| 7a | Data extraction | 0.12 | -0.05 | 0.30 | | 0.10 | -0.08 | 0.28 | | 0.52 | 0.13 | 0.91 | |
| 7b | Description of studies | 0.34 | 0.06 | 0.62 | | 0.39 | 0.09 | 0.68 | | 0.57 | 0.37 | 0.77 | |
| ^7c | PF effect calculations | 0.12 | -0.01 | 0.25 | | 0.11 | -0.10 | 0.33 | | 0.32 | 0.13 | 0.50 | |
| 8a | RoB process | 0.27 | 0.02 | 0.51 | | 0.34 | 0.06 | 0.63 | | 0.71 | 0.41 | 1.00 | |
| 8b | RoB technique | 0.35 | 0.16 | 0.53 | | 0.35 | 0.20 | 0.50 | | 0.59 | 0.15 | 1.02 | |
| ^9a | Synthesis interpretability | 0.31 | -0.02 | 0.64 | | 0.24 | -0.11 | 0.59 | | 0.54 | 0.27 | 0.81 | |
| ^9b | Meta-Analysis | 0.44 | 0.08 | 0.80 | | 0.57 | 0.24 | 0.91 | | 0.62 | 0.42 | 0.82 | |
| ^10 | Small study effects | 0.22 | -0.05 | 0.49 | | 0.21 | -0.12 | 0.55 | | 0.53 | 0.39 | 0.67 | |
| 11 | Impact of RoB | 0.25 | -0.03 | 0.54 | | 0.27 | -0.07 | 0.62 | | 0.59 | 0.32 | 0.87 | |
| ^12 | Heterogeneity | 0.16 | 0.01 | 0.31 | | 0.21 | 0.06 | 0.36 | | 0.45 | 0.31 | 0.60 | |
| 13 | Conflicts of interest | 0.30 | -0.03 | 0.64 | | 0.24 | -0.11 | 0.59 | | 0.48 | 0.16 | 0.80 | |
| 14 | Certainty of findings | 0.34 | 0.06 | 0.62 | | 0.38 | 0.05 | 0.71 | | 0.46 | 0.27 | 0.65 | |
| Final Appraisal | | 0.45 | 0.10 | 0.80 | | 0.47 | 0.04 | 0.90 | | 0.65 | 0.43 | 0.86 | |

**Table A: Kappa scores for Interrater, Inter-pair, and Intrapair agreement, with all answering options.** Fleiss' Kappa was used for Interrater and Inter-pair agreement calculations, and average Cohen's Kappa is displayed for intrapair agreement. Benchmark interpretation per Landis and Koch's benchmark categories, < 0, Poor; 0.0-0.2, Slight; 0.2-0.4, Fair; 0.4-0.6, Moderate; 0.6-0.8, Substantial; and 0.8-1.0, Almost Perfect. Confidence intervals were capped at 1.00

C Kap, average Cohen's Kappa; F Kap, Fleiss' Kappa; I, interpretation; 95%CI, 95% Confidence Interval.

| Question | | Interrater | | | | Inter-pair | | | | Intrapair | | | |
|---|---|---|---|---|---|---|---|---|---|---|---|---|---|
| | | F Kap | 95%CI | | I | F Kap | 95%CI | | I | C Kap | 95%CI | | I |
| 1 | Research Question | 0.27 | -0.02 | 0.56 | | 0.34 | 0.08 | 0.61 | | 0.87 | 0.65 | 1.00 | |
| 2a | Protocol Registration | 0.72 | 0.15 | 1.00 | | 0.68 | 0.04 | 1.00 | | 0.85 | 0.62 | 1.00 | |
| ^2b | Deviations from protocol | 0.34 | -0.01 | 0.69 | | 0.40 | -0.02 | 0.82 | | 0.53 | 0.23 | 0.83 | |
| 3 | Included study designs | 0.05 | -0.03 | 0.14 | | -0.04 | -0.10 | 0.02 | | 0.54 | 0.01 | 1.00 | |
| 4 | Search strategy | 0.03 | -0.05 | 0.11 | | -0.04 | -0.10 | 0.02 | | 0.33 | -0.10 | 0.77 | |
| 5 | Inclusion process | 0.00 | -0.06 | 0.05 | | 0.14 | 0.06 | 0.21 | | 0.39 | -0.14 | 0.92 | |
| 6 | Excluded studies | 0.86 | 0.66 | 1.00 | | 0.90 | 0.71 | 1.00 | | 0.96 | 0.86 | 1.00 | |
| 7a | Data extraction | -0.02 | -0.05 | 0.01 | | -0.02 | -0.06 | 0.03 | | 0.71 | 0.26 | 1.00 | |
| 7b | Description of studies | 0.11 | -0.02 | 0.24 | | 0.11 | -0.08 | 0.30 | | 0.53 | 0.06 | 1.00 | |
| ^7c | PF effect calculations | 0.22 | -0.04 | 0.48 | | 0.21 | -0.13 | 0.55 | | 0.49 | 0.24 | 0.75 | |
| 8a | RoB process | 0.36 | 0.21 | 0.51 | | 0.46 | 0.36 | 0.56 | | 0.71 | 0.26 | 1.00 | |
| 8b | RoB technique | 0.21 | 0.09 | 0.34 | | 0.14 | 0.06 | 0.21 | | 0.71 | 0.26 | 1.00 | |
| ^9a | Synthesis interpretability | 0.44 | -0.02 | 0.89 | | 0.36 | -0.14 | 0.86 | | 0.63 | 0.35 | 0.91 | |
| ^9b | Meta-Analysis | 0.72 | 0.37 | 1.00 | | 0.91 | 0.74 | 1.00 | | 0.81 | 0.57 | 1.00 | |
| ^10 | Small study effects | 0.30 | -0.02 | 0.62 | | 0.20 | -0.19 | 0.60 | | 0.66 | 0.38 | 0.95 | |
| 11 | Impact of RoB | 0.23 | -0.02 | 0.49 | | 0.18 | -0.07 | 0.43 | | 0.59 | 0.25 | 0.92 | |
| ^12 | Heterogeneity | 0.43 | 0.23 | 0.63 | | 0.56 | 0.39 | 0.73 | | 0.57 | 0.26 | 0.87 | |
| 13 | Conflicts of interest | 0.08 | -0.05 | 0.21 | | 0.05 | -0.09 | 0.18 | | 0.51 | 0.04 | 0.98 | |
| 14 | Certainty of findings | 0.26 | 0.00 | 0.52 | | 0.30 | -0.03 | 0.63 | | 0.45 | 0.15 | 0.76 | |

**Table B: Kappa scores for Interrater, Inter-pair, and Intrapair agreement, with collapsed answering options.** Fleiss' Kappa was used for Interrater and Inter-pair agreement calculations, and average Cohen's Kappa is displayed for intrapair agreement. Benchmark interpretation per Landis and Koch's benchmark categories, < 0, Poor; 0.0-0.2, Slight; 0.2-0.4, Fair; 0.4-0.6, Moderate; 0.6-0.8, Substantial; and 0.8-1.0, Almost Perfect. Confidence intervals were capped at 1.00

C Kap, average Cohen's Kappa; F Kap, Fleiss' Kappa; I, interpretation; 95%CI, 95% Confidence Interval.
